## Supplementary Information. for "Population clustering of structural brain aging and its association with brain development"

#### Contents


**Appendix 1: supplementary methods**

**Appendix 1: supplementary tables**

**Appendix 1—table 1.** ICD-10 primary and secondary diagnostic codes for exclusion criteria.

**Appendix 1—table 2.** Self-reported illness codes for exclusion criteria.

**Appendix 1—table 3.** Cortical and subcortical brain regions.

**Appendix 1—table 4.** Loadings matrix for the first 15 principal components.

**Appendix 1—table 5.** Baseline and demographic characteristics for participants in the total population and stratified by brain aging patterns.

**Appendix 1—table 6.** Associations between results of biological aging biomarkers and subgroups stratified by whole-brain TGMV trajectories.

**Appendix 1—table 7.** Associations between results of cognitive function tests and subgroups stratified by whole-brain TGMV trajectories.

**Appendix 1—table 8.** Genome-wide association study details.

**Appendix 1—table 9.** Polygenic Risk Scores comparisons between two subgroups. Data supporting these scores were obtained either entirely from external GWAS data (the Standard PRS set).

**Appendix 1—table 10.** Polygenic Risk Scores comparisons between two subgroups. Data supporting these scores were obtained external and internal UK Biobank data (the Enhanced PRS set).

**Appendix 1—table 11.** Most significant single-variant associations ($P < 5 \times 10^{-8}$) detected in the GWAS analyses.

**Appendix 1—table 12.** Association between gene expression profiles of mapped genes and estimated APC during brain development.

**Appendix 1—table 13.** Association between gene expression profiles of mapped genes and estimated APC during brain aging.

**Appendix 1—table 14.** Model evaluation results using relative measures: AIC, BIC, likelihood ratio test and intra-class correlation (ICC).

**Appendix 1: supplementary figures**

**Appendix 1—figure 1.** The sample selection workflow.

**Appendix 1—figure 2.** Estimated rates of change in regional volumes for 33 bilateral brain regions.

**Appendix 1—figure 3.** Stratification of the identified brain aging patterns using linear and non-linear dimensionality reduction methods.

**Appendix 1—figure 4** Effect size for comparing each individual blood biochemical metric (used to calculate the PhenoAge) between participants with brain aging patterns 1 and 2.

**Appendix 1—figure 5.** Gene set enrichment of Kyoto Encyclopedia of Genes and Genomes (KEGG) pathways and gene ontology (GO) of biological processes.

**Appendix 1—figure 6.** Optimal number of clusters was chosen using elbow method (a) and silhouette method (b).

**Appendix 1: Supplementary Methods**

**Cognitive assessment**

**Reaction time** This cognitive function test is based on 12 rounds of the card-game 'Snap'. The participant is shown two cards at a time; if both cards are the same, they press a button-box that is on the table in front of them as quickly as possible. The score used for analysis is mean time to correctly identify matches (UK Biobank data field 20023), which is the mean duration to first press of snap-button summed over rounds in which both cards matched.

**Numeric memory** The participant was shown a 2-digit number to remember. The number then disappeared and after a short while they were asked to enter the number onto the screen. The number became one digit longer each time they remembered correctly (up to a maximum of 12 digits). This test is available for a subset of participants. The score used for analysis is maximum digits remembered correctly (UK Biobank data field 4282), which is longest number correctly recalled during the numeric memory test.

**Fluid intelligence / reasoning** 'Fluid intelligence' is defined as the capacity to solve problems that require logic and reasoning ability, independent of acquired knowledge. The participant has 2 minutes to complete as many questions as possible from the test. This test was incorporated into the touchscreen towards the end of recruitment. The score used for analysis is fluid intelligence score (UK Biobank data field 20016), which is a simple unweighted sum of the number of correct answers given to the 13 fluid intelligence questions. Participants who did not answer all of the questions within the allotted 2 minutes limit are scored as zero for each of the unattempted questions.

**Trail making** The participant was presented with sets of digits/letters in circles scattered around the screen and asked to click on them sequentially according to a specific algorithm. The scores used for analysis are duration to complete numeric path (trail #1) (UK Biobank data field 6348) and duration to complete alphanumeric path (trail #2) (UK Biobank data field 6350).

**Matrix pattern completion** The participant was presented with a series of matrix pattern blocks with an element missing and asked to select the element that best completed the pattern from a range of displayed choices. The score used for analysis is number of puzzles correctly solved (UK Biobank data field 6373).

**Symbol digit substitution** The participant was presented with one grid linking symbols to single-digit integers and a second grid containing only the symbols. They were then asked to indicate the numbers attached to each of the symbols in the second grid using the first one as a key. The score used for analysis is number of symbol digit matches made correctly (UK Biobank data field 23324).

**Tower rearranging** The participant was presented with an illustration of three pegs (towers) on which three differently-colored hoops had been placed. The were then asked to indicate how many moves it would take to re-arrange the hoops into another specific position. The score used for analysis is number of puzzles correct (UK Biobank data field 21004).

**Paired associate learning** In the paired associate learning test the participants were shown 12 pairs of words (for 30 seconds in total) then, after an interval (in which they did a different test), presented with the first word of 10 of these pairs and asked to select the matching second word from a choice of 4 alternatives. The words were presented in the order: huge, happy, tattered, old, long, red, sulking, pretty, tiny and new. The score used for analysis is number of word pairs correctly associated (UK Biobank data field 21097), which is the number of word pairs correctly associated out of ten attempts.

**Prospective memory** Early in the touchscreen cognitive section, the participant is shown the message "At the end of the games we will show you four colored shapes and ask you to touch the Blue Square. However, to test your memory, we want you to actually touch the Orange Circle instead." The score used for analysis is prospective memory result (UK Biobank data field 20018), which condenses the results of the prospective memory test into 3 groups ("0" : instruction not recalled, either skipped or incorrect, "1" : correct recall on first attempt, "2" : correct recall on second attempt). We divided the test results into two groups for simplicity of analysis: "0" indicating no correct recall, and the combination of "1" and "2" indicating correct recall.

**Pairs matching** Participants are asked to memorize the position of as many matching pairs of cards as possible. The cards are then turned face down on the screen and the participant is asked to touch as many pairs as possible in the fewest tries. Multiple rounds were conducted. The first round used 3 pairs of cards and the second 6 pairs of cards. In the pilot phase an additional (i.e., third) round was conducted using 6 pairs of cards. However this was dropped from the main study as the extra set of results were very similar to the second and not felt to add significant new information. The score used for analysis is number of incorrect matches in round 2 (UK Biobank data field 399.2).

### Appendix 1: Supplementary Tables

#### Appendix 1—table 1. ICD-10 primary and secondary diagnostic codes for exclusion criteria.

| Condition | Code |
|---|---|
| Malignant neoplasm | C70, C71 |
| Dementia | F00, F01, F02, F03, F04 |
| Mental and behavioural disorders due to psychoactive substance use | F10-F19 |
| Schizophrenia, schizotypal and delusional disorders | F20-F29 |
| Mood [affective] disorders | F30-F39 |
| Mental retardation | F70-F79 |
| Disorders of psychological development | F80-F89 |
| Hyperkinetic disorders | F90 |
| Inflammatory diseases of the central nervous system | G00-G09 |
| Systemic atrophies primarily affecting the central nervous system | G10, G11, G122, G13 |
| Extrapyramidal and movement disorders | G20, G21, G22, G23 |
| Other degenerative diseases of the nervous system | G30-G32 |
| Demyelinating diseases of the central nervous system | G35-G37 |
| Episodic and paroxysmal disorders | G40, G41, G45, G46 |
| Infantile cerebral palsy | G80 |
| Cerebrovascular diseases | I60-I69 |
| Down's syndrome | Q90 |
| Intracranial injury | S06 |

**Appendix 1—table 2. Self-reported illness codes for exclusion criteria.**

| Field ID | Condition | Code |
|---|---|---|
| 20001 | Brain cancer/primary malignant brain tumour | 1032 |
| | Meningeal cancer/malignant meningioma | 1031 |
| 20002 | Benign neuroma | 1683 |
| | Brain abscess/intracranial abscess | 1245 |
| | Brain haemorrhage | 1491 |
| | Cerebral aneurysm | 1425 |
| | Cerebral palsy | 1433 |
| | Chronic/degenerative neurological problem | 1258 |
| | dementia/alzheimers/cognitive impairment | 1263 |
| | Encephalitis | 1246 |
| | Epilepsy | 1264 |
| | Fracture skull/head | 1626 |
| | Head injury | 1266 |
| | Ischaemic stroke | 1583 |
| | Meningioma/benign meningeal tumour | 1659 |
| | Meningitis | 1247 |
| | Motor Neurone Disease | 1259 |
| | Multiple Sclerosis | 1261 |
| | Nervous system infection | 1244 |
| | Neurological injury/trauma | 1240 |
| | Other demyelinating disease (not Multiple Sclerosis) | 1397 |
| | Other neurological problem | 1434 |
| | Parkinson's Disease | 1262 |
| | Spina Bifida | 1524 |
| | Stroke | 1081 |
| | Subarachnoid haemorrhage | 1086 |
| | Subdural haemorrhage/haematoma | 1083 |
| | Transient ischaemic attack | 1082 |

**Appendix 1—table 3. Cortical and subcortical brain regions.**

| **Desikan–Killiany Atlas** |
|---|
| bankssts |
| caudal anterior cingulate |
| caudal middle frontal |
| cuneus |
| entorhinal |
| fusiform |
| inferior parietal |
| inferior temporal |
| isthmus cingulate |
| lateral occipital |
| lateral orbitofrontal |
| lingual |
| medial orbitofrontal |
| middle temporal |
| parahippocampal |
| paracentral |
| pars opercularis |
| pars orbitalis |
| pars triangularis |
| pericalcarine |
| postcentral |
| posterior cingulate |
| precentral |
| precuneus |
| rostral anterior cingulate |
| rostral middle frontal |
| superior frontal |
| superior parietal |
| superior temporal |
| supramarginal |
| frontal pole |
| transverse temporal |
| insula |
| **ASEG Atlas** |
| thalamus proper |
| caudate |
| putamen |
| pallidum |
| hippocampus |
| amygdala |
| accumbens area |

#### Appendix 1—table 4. Loadings matrix for the first 15 principal components.

| | PC1 | PC2 | PC3 | PC4 | PC5 | PC6 | PC7 | PC8 | PC9 | PC10 | PC11 | PC12 | PC13 | PC14 | PC15 |
|---|---|---|---|---|---|---|---|---|---|---|---|---|---|---|---|
| **Cortical** | | | | | | | | | | | | | | | |
| bankssts | 0.12 | -0.25 | 0.01 | 0.30 | -0.13 | 0.01 | 0.09 | 0.34 | 0.04 | 0.05 | -0.10 | 0.16 | -0.09 | -0.20 | 0.02 |
| caudal.anterior.cingulate | 0.12 | -0.06 | -0.02 | 0.13 | 0.39 | 0.28 | 0.13 | -0.09 | -0.37 | 0.24 | 0.05 | 0.03 | -0.04 | -0.13 | 0.07 |
| caudal.middle.frontal | 0.15 | 0.01 | -0.10 | -0.11 | 0.34 | -0.21 | 0.10 | 0.06 | 0.03 | -0.28 | -0.23 | -0.20 | 0.24 | 0.00 | 0.25 |
| cuneus | 0.13 | 0.47 | -0.01 | 0.06 | -0.01 | 0.00 | 0.07 | 0.12 | 0.01 | 0.07 | 0.12 | -0.05 | -0.09 | 0.08 | -0.07 |
| entorhinal | 0.09 | 0.07 | 0.13 | 0.08 | 0.10 | 0.17 | -0.53 | -0.18 | 0.06 | 0.04 | -0.19 | -0.10 | 0.13 | 0.01 | -0.04 |
| fusiform | 0.18 | -0.03 | 0.01 | 0.13 | -0.02 | 0.02 | -0.20 | -0.02 | 0.15 | 0.30 | -0.01 | -0.28 | -0.23 | 0.08 | 0.21 |
| inferior.parietal | 0.15 | -0.18 | 0.01 | 0.39 | -0.09 | 0.01 | 0.14 | -0.04 | 0.16 | 0.05 | -0.20 | -0.02 | -0.15 | -0.02 | 0.02 |
| inferior.temporal | 0.16 | -0.13 | 0.01 | 0.26 | -0.01 | 0.15 | -0.04 | -0.10 | 0.14 | -0.08 | -0.08 | -0.25 | 0.20 | 0.29 | -0.01 |
| isthmus.cingulate | 0.15 | 0.23 | 0.02 | 0.11 | -0.14 | -0.03 | 0.08 | -0.03 | -0.23 | -0.07 | -0.22 | 0.16 | 0.43 | 0.04 | 0.11 |
| lateral.occipital | 0.16 | 0.30 | 0.00 | 0.12 | -0.02 | 0.02 | 0.05 | 0.10 | 0.16 | 0.18 | 0.06 | -0.23 | -0.23 | 0.12 | 0.17 |
| lateral.orbitofrontal | 0.22 | -0.04 | -0.08 | -0.17 | -0.05 | 0.15 | -0.02 | -0.15 | 0.09 | -0.02 | -0.01 | 0.40 | -0.01 | 0.22 | 0.17 |
| lingual | 0.12 | 0.43 | 0.03 | 0.14 | -0.01 | -0.02 | 0.07 | 0.11 | -0.01 | -0.03 | -0.10 | 0.16 | 0.02 | -0.09 | -0.08 |
| medial.orbitofrontal | 0.21 | -0.06 | -0.06 | -0.06 | -0.05 | 0.10 | -0.01 | -0.13 | 0.18 | -0.06 | 0.13 | 0.17 | 0.05 | 0.01 | -0.25 |
| middle.temporal | 0.17 | -0.17 | -0.01 | 0.27 | -0.14 | 0.10 | 0.21 | 0.25 | 0.09 | -0.15 | -0.09 | 0.04 | 0.07 | 0.05 | -0.11 |
| parahippocampal | 0.12 | 0.09 | 0.13 | 0.05 | 0.06 | 0.14 | -0.46 | 0.02 | -0.02 | 0.09 | -0.22 | 0.09 | 0.03 | -0.32 | 0.02 |
| paracentral | 0.19 | -0.05 | -0.10 | -0.07 | 0.18 | -0.23 | -0.11 | -0.06 | 0.01 | 0.03 | -0.02 | 0.06 | -0.35 | -0.01 | -0.35 |
| pars.opercularis | 0.16 | -0.02 | -0.06 | -0.23 | -0.19 | 0.10 | 0.14 | 0.01 | -0.16 | -0.13 | -0.43 | -0.29 | -0.16 | -0.06 | 0.00 |
| pars.orbitalis | 0.18 | 0.02 | -0.05 | -0.17 | -0.11 | 0.24 | 0.03 | -0.17 | 0.23 | 0.08 | 0.01 | 0.41 | -0.14 | 0.05 | 0.24 |
| pars.triangularis | 0.16 | 0.02 | -0.03 | -0.35 | -0.26 | 0.23 | 0.13 | -0.05 | -0.06 | -0.06 | -0.25 | -0.17 | -0.21 | -0.08 | -0.05 |
| pericalcarine | 0.10 | 0.47 | -0.01 | 0.08 | -0.01 | 0.01 | 0.09 | 0.13 | -0.02 | -0.01 | 0.02 | 0.08 | -0.03 | 0.03 | -0.17 |
| postcentral | 0.20 | -0.04 | -0.11 | 0.06 | 0.16 | -0.29 | -0.09 | 0.12 | 0.08 | 0.03 | -0.09 | 0.10 | 0.04 | 0.01 | 0.04 |
| posterior.cingulate | 0.18 | -0.08 | -0.04 | 0.02 | 0.24 | 0.09 | 0.08 | -0.02 | -0.30 | 0.01 | -0.07 | 0.10 | -0.05 | -0.12 | -0.31 |
| precentral | 0.21 | -0.01 | -0.10 | -0.08 | 0.28 | -0.29 | -0.09 | 0.08 | 0.15 | -0.05 | -0.15 | 0.13 | -0.06 | 0.05 | 0.12 |
| precuneus | 0.20 | 0.02 | -0.02 | 0.13 | -0.24 | -0.26 | 0.01 | -0.35 | -0.20 | -0.04 | 0.06 | -0.04 | 0.00 | 0.03 | 0.07 |
| rostral.anterior.cingulate | 0.15 | -0.07 | -0.07 | 0.05 | 0.31 | 0.21 | 0.15 | -0.05 | -0.30 | 0.13 | 0.11 | -0.06 | 0.00 | 0.18 | 0.15 |
| rostral.middle.frontal | 0.20 | 0.02 | -0.05 | -0.05 | 0.10 | 0.14 | 0.22 | -0.11 | 0.17 | -0.11 | 0.24 | -0.14 | 0.19 | -0.22 | 0.14 |
| superior.frontal | 0.22 | -0.03 | -0.09 | -0.21 | 0.14 | -0.13 | -0.01 | 0.06 | 0.20 | -0.18 | 0.06 | -0.11 | 0.02 | 0.00 | -0.16 |
| superior.parietal | 0.17 | 0.02 | -0.05 | 0.14 | -0.18 | -0.29 | 0.01 | -0.45 | -0.16 | 0.00 | 0.14 | -0.08 | -0.08 | -0.06 | 0.04 |
| superior.temporal | 0.21 | -0.13 | -0.02 | -0.06 | -0.10 | 0.01 | -0.07 | 0.40 | -0.07 | 0.13 | 0.24 | -0.05 | 0.02 | -0.06 | 0.08 |
| supramarginal | 0.17 | -0.15 | -0.08 | 0.06 | -0.24 | -0.23 | -0.13 | 0.03 | -0.21 | 0.04 | 0.11 | 0.10 | 0.14 | -0.08 | -0.06 |
| frontal.pole | 0.16 | 0.03 | -0.10 | 0.01 | 0.00 | 0.16 | 0.03 | -0.13 | 0.28 | 0.05 | 0.28 | -0.16 | 0.26 | -0.38 | -0.22 |
| transverse.temporal | 0.15 | -0.01 | -0.10 | -0.27 | -0.17 | -0.10 | -0.16 | 0.25 | -0.22 | 0.19 | 0.24 | -0.10 | 0.09 | 0.04 | 0.16 |
| insula | 0.19 | -0.08 | 0.01 | -0.22 | -0.12 | 0.10 | -0.07 | 0.11 | -0.06 | 0.15 | -0.08 | 0.00 | 0.19 | 0.16 | -0.23 |
| **Subcortical** | | | | | | | | | | | | | | | |
| thalamus.proper | 0.09 | -0.02 | 0.31 | -0.08 | 0.03 | -0.08 | 0.08 | 0.00 | 0.02 | -0.21 | 0.11 | 0.10 | -0.19 | -0.37 | 0.33 |
| caudate | 0.06 | -0.03 | 0.32 | -0.12 | 0.02 | -0.10 | 0.15 | -0.02 | 0.11 | 0.31 | -0.11 | 0.09 | 0.27 | 0.14 | 0.12 |
| putamen | 0.08 | -0.03 | 0.42 | -0.12 | 0.03 | -0.13 | 0.13 | -0.03 | 0.05 | 0.22 | -0.05 | -0.14 | 0.07 | -0.02 | -0.17 |
| pallidum | 0.03 | -0.03 | 0.42 | -0.07 | 0.00 | -0.13 | 0.16 | -0.06 | 0.04 | 0.20 | -0.07 | 0.00 | 0.04 | -0.20 | -0.06 |
| hippocampus | 0.12 | 0.01 | 0.33 | 0.04 | -0.02 | 0.11 | -0.20 | 0.08 | -0.16 | -0.39 | 0.09 | 0.03 | -0.11 | -0.03 | 0.05 |
| amygdala | 0.13 | -0.03 | 0.31 | 0.05 | -0.01 | 0.12 | -0.13 | 0.09 | -0.07 | -0.35 | 0.25 | -0.09 | -0.02 | 0.24 | -0.01 |
| accumbens.area | 0.11 | -0.05 | 0.31 | -0.02 | 0.12 | -0.07 | 0.11 | -0.04 | 0.00 | -0.03 | 0.10 | 0.09 | -0.14 | 0.34 | -0.18 |

**Appendix 1—table 5. Baseline and demographic characteristics for participants in the total population and stratified by brain aging patterns.**

| | Total (n=37,013) | Pattern 1 (n=18,929) | Pattern 2 (n=18,084) |
|---|---|---|---|
| **Age (years), mean (SD)** | 63.9 (7.63) | 63.9 (7.64) | 63.8 (7.63) |
| **Female, n (%)** | 19,958 (53.9) | 10,117 (53.4) | 9,841 (54.4) |
| **Ethnicity, n (%)** | | | |
| **White** | 34,219 (92.5) | 17,509 (92.5) | 16,710 (92.4) |
| **Mixed** | 1,137 (3.1) | 573 (3.0) | 564 (3.1) |
| **Asian or Asian British** | 1,210 (3.3) | 612 (3.2) | 598 (3.3) |
| **Other** | 447 (1.2) | 235 (1.2) | 212 (1.2) |
| **Smoking status, n (%)** [a] | | | |
| **Never smoker** | 23,633 (64.4) | 12,269 (65.4) | 11,364 (63.4) |
| **Previous smoker** | 12,213 (33.3) | 6,085 (32.4) | 6,128 (34.2) |
| **Current smoker** | 833 (2.3) | 414 (2.2) | 419 (2.3) |
| **TDI, mean (SD)** [b] | -1.94 (2.69) | -1.97 (2.66) | -1.90 (2.71) |
| **BMI (kg/m^2), mean (SD)** [c] | 26.4 (4.32) | 26.3 (4.17) | 26.5 (4.46) |
| **Years of Schooling, mean (SD)** [d] | 16.8 (4.32) | 16.9 (4.29) | 16.8 (4,35) |

Abbreviations: TDI = Townsend Deprivation Index, BMI = Body Mass Index.

[a] Missing 334

[b] Missing 36

[c] Missing 1,937

[d] Missing 337

**Appendix 1—table 6. Associations between results of biological aging biomarkers and subgroups stratified by whole-brain TGMV trajectories.** Cohen's d measures the standardized difference of means between brain aging pattern 2 and brain aging pattern 1.

| Biological aging biomarkers | Brain aging pattern 1 | | Brain aging pattern 2 | |
|---|---|---|---|---|
| | N | Mean(SD) | N | Mean(SD) |
| LTL | 17,691 | 0.083(0.98) | 16,876 | 0.055(0.97) |
| PhenoAge | 15,228 | 41.35(8.17) | 14,323 | 41.58(8.32) |

| Biological aging biomarkers | Unadjusted | | | Adjusted | | |
|---|---|---|---|---|---|---|
| | Cohen's d (95% CI) | P | P.Bonferroni | Cohen's d (95% CI) | P | P.Bonferroni |
| LTL | -0.028 (-0.049, -0.007) | 0.009 | 0.019 | -0.030 (-0.051, -0.009) | 0.006 | 0.011 |
| PhenoAge | 0.027 (0.004, 0.050) | 0.019 | 0.039 | 0.092 (0.070, 0.116) | 3.05E-15 | 6.11E-15 |

**Appendix 1—table 7. Associations between results of cognitive function tests and subgroups stratified by whole-brain TGMV trajectories.** Cohen's d measures the standardized difference of means between brain aging pattern 2 and brain aging pattern 1.

| Cognitive functions | Brain aging pattern 1 | | Brain aging pattern 2 | |
|---|---|---|---|---|
| | N | Mean(SD) | N | Mean(SD) |
| **Reaction time** | 17,749 | -594.70(108.15) | 16,831 | -594.68(110.50) |
| **Numeric memory** | 13,350 | 6.82(1.26) | 12,346 | 6.72(1.27) |
| **Fluid intelligence** | 17,580 | 6.73(2.06) | 16,612 | 6.53(2.04) |
| **Trail making A** | 13,052 | -224.50(84.82) | 12,036 | -227.56(84.85) |
| **Trail making B** | 12,743 | -563.06(246.74) | 11,753 | -576.55(260.75) |
| **Matrix pattern completion** | 13,064 | 8.07(2.11) | 12,064 | 7.91(2.14) |
| **Symbol digit substitution** | 13,077 | 19.08(5.15) | 12,056 | 18.87(5.35) |
| **Tower rearranging** | 12,952 | 9.96(3.20) | 11,958 | 9.83(3.23) |
| **Paired associate learning** | 13,184 | 7.01(2.59) | 12,198 | 6.88(2.65) |
| **Prospective memory** | 17,831 | N/A | 16,949 | N/A |
| **Pairs matching** | 17,840 | -3.58(2.90) | 16,956 | -3.67(2.94) |

| Cognitive functions | Unadjusted | | | Adjusted | | |
|---|---|---|---|---|---|---|
| | Cohen's d (95% CI) | P | P.FDR | Cohen's d (95% CI) | P | P.FDR |
| **Reaction time** | 0.000 (-0.021, 0.021) | 0.99 | 0.99 | 0.006 (-0.016, 0.028) | 0.61 | 0.61 |
| **Numeric memory** | -0.082 (-0.106, -0.057) | 5.97E-11 | 3.28E-10 | -0.080 (-0.106, -0.055) | 8.99E-10 | 4.95E-09 |
| **Fluid intelligence** | -0.102 (-0.123, -0.080) | 5.94E-21 | 6.54E-20 | -0.99 (-0.121, -0.077) | 3.30E-18 | 3.63E-17 |
| **Trail making A** | -0.036 (-0.061, -0.011) | 0.004 | 0.006 | -0.050 (-0.074, -0.024) | 1.46E-04 | 2.29E-04 |
| **Trail making B** | -0.053 (-0.078, -0.028) | 3.16E-05 | 8.68E-05 | -0.067 (-0.093, -0.041) | 6.16E-07 | 1.69E-06 |
| **Matrix pattern completion** | -0.076 (-0.101, -0.051) | 1.84E-09 | 6.74E-09 | -0.078 (-0.104, -0.052) | 3.67E-09 | 1.35E-08 |
| **Symbol digit substitution** | -0.040 (-0.065, -0.015) | 0.002 | 0.002 | -0.053 (-0.079, -0.027) | 6.15E-05 | 1.13E-04 |
| **Tower rearranging** | -0.041 (-0.066, -0.016) | 0.001 | 0.002 | -0.049 (-0.075, -0.023) | 2.18E-04 | 3.00E-04 |
| **Paired associate learning** | -0.051 (-0.076, -0.027) | 4.64E-05 | 1.02E-04 | -0.054 (-0.079, -0.028) | 4.89E-05 | 1.08E-04 |
| **Prospective memory** | OR: 0.943 (0.891, 0.999) | 0.047 | 0.052 | OR: 0.940 (0.883, 1.000) | 0.052 | 0.057 |
| **Pairs matching** | -0.029 (-0.050, -0.008) | 0.006 | 0.008 | -0.033 (-0.055, -0.011) | 0.003 | 0.004 |

**Appendix 1—table 8. Genome-wide association study details.** Loci associated with risk were thresholded at $P < 5 \times 10^{-8}$, then distance-based clumping was used to define independently significant loci.

| Study | Number cases | Number controls | Number of genome-wide independently significant loci | Download link |
|---|---|---|---|---|
| Attention deficit hyperactivity disorder[1] | 20,183 | 35,191 | 12 | https://figshare.com/ndownloader/files/28169253 |
| Autism spectrum disorder[2] | 18,381 | 27,969 | 5 | https://figshare.com/ndownloader/files/28169292 |
| Alzheimer's disease[3] | 71,880 | 383,378 | 25 | https://ctg.cncr.nl/software/summary_statistics |
| Parkinson's disease[4] | 37,688, and 18,618 (proxy-cases) | 1,417,791 | 90 | https://drive.google.com/file/d/1FZ9UL99LAqyWnyNBxxlx6qOUlfAnublN/view?usp=sharing |
| Bipolar disorder[5] | 41,917 | 371,549 | 64 | https://figshare.com/ndownloader/files/40036705 |
| Major depressive disorder[6] | 135,458 | 344,901 | 44 | https://figshare.com/ndownloader/files/39504667 |
| Schizophrenia[7] | 76,755 | 243,649 | 287 | https://figshare.com/ndownloader/files/34517828 |
| Delayed Brain Development[8] | 7,662 (proxy phenotype, continuous) | | 1 | https://delayedneurodevelopment.page.link/amTC |

**Appendix 1—table 9. Polygenic Risk Scores comparisons between two subgroups. Data supporting these scores were obtained either entirely from external GWAS data (the Standard PRS set).** The bold P values reflect significance after FDR correction.

| Trait | n1 | n2 | statistic | p | p.adjust |
|---|---|---|---|---|---|
| AAM | 18,429 | 17,586 | -2.218 | 0.027 | 0.080 |
| AMD | 18,429 | 17,586 | 1.753 | 0.080 | 0.169 |
| AD | 18,429 | 17,586 | 0.735 | 0.462 | 0.616 |
| AST | 18,429 | 17,586 | -0.861 | 0.389 | 0.543 |
| AF | 18,429 | 17,586 | -0.100 | 0.920 | 0.945 |
| BD | 18,429 | 17,586 | 3.557 | 3.75E-04 | **0.002** |
| BMI | 18,429 | 17,586 | -3.309 | 0.001 | **0.005** |
| CRC | 18,429 | 17,586 | -0.544 | 0.586 | 0.703 |
| BC | 18,429 | 17,586 | -3.140 | 0.002 | **0.008** |
| CVD | 18,429 | 17,586 | -2.104 | 0.035 | 0.091 |
| CED | 18,429 | 17,586 | 1.046 | 0.296 | 0.484 |
| CAD | 18,429 | 17,586 | -1.588 | 0.112 | 0.202 |
| CD | 18,429 | 17,586 | -0.094 | 0.925 | 0.945 |
| EOC | 18,429 | 17,586 | -2.183 | 0.029 | 0.080 |
| EBMDT | 18,429 | 17,586 | -11.343 | <1.00E-20 | **<1.00E-20** |
| HBA1C_DF | 18,429 | 17,586 | -2.948 | 0.003 | **0.013** |
| HEIGHT | 18,429 | 17,586 | 6.658 | 2.81E-11 | **3.37E-10** |
| HDL | 18,429 | 17,586 | 0.884 | 0.377 | 0.543 |
| HT | 18,429 | 17,586 | -3.539 | 4.02E-04 | **0.002** |
| IOP | 18,429 | 17,586 | -1.605 | 0.109 | 0.202 |
| ISS | 18,429 | 17,586 | -2.383 | 0.017 | 0.056 |
| LDL_SF | 18,429 | 17,586 | -0.686 | 0.492 | 0.627 |
| MEL | 18,429 | 17,586 | 2.025 | 0.043 | 0.103 |
| MS | 18,429 | 17,586 | -0.069 | 0.945 | 0.945 |
| OP | 18,429 | 17,586 | 12.029 | <1.00E-20 | **<1.00E-20** |
| PD | 18,429 | 17,586 | 1.456 | 0.145 | 0.249 |
| POAG | 18,429 | 17,586 | -0.856 | 0.392 | 0.543 |
| PC | 18,429 | 17,586 | -0.240 | 0.810 | 0.941 |
| PSO | 18,429 | 17,586 | -1.781 | 0.075 | 0.169 |
| RA | 18,429 | 17,586 | -2.437 | 0.015 | 0.053 |
| SCZ | 18,429 | 17,586 | 0.158 | 0.874 | 0.945 |
| SLE | 18,429 | 17,586 | 1.695 | 0.090 | 0.180 |
| T1D | 18,429 | 17,586 | 0.666 | 0.505 | 0.627 |
| T2D | 18,429 | 17,586 | -5.523 | 3.35E-08 | **3.02E-07** |
| UC | 18,429 | 17,586 | 0.883 | 0.377 | 0.543 |
| VTE | 18,429 | 17,586 | -0.170 | 0.865 | 0.945 |

**Appendix 1—table 10. Polygenic Risk Scores comparisons between two subgroups. Data supporting these scores were obtained external and internal UK Biobank data (the Enhanced PRS set).** The bold P values reflect significance after FDR correction.

| Trait | n1 | n2 | statistic | p | p.adjust |
|---|---|---|---|---|---|
| AAM | 3,407 | 3,409 | -1.708 | 0.088 | 0.344 |
| AMD | 3,407 | 3,409 | 0.547 | 0.584 | 0.931 |
| AD | 3,407 | 3,409 | 0.756 | 0.450 | 0.820 |
| APOEA | 3,407 | 3,409 | 0.023 | 0.982 | 0.993 |
| APOEB | 3,407 | 3,409 | 0.119 | 0.905 | 0.968 |
| AST | 3,407 | 3,409 | 0.112 | 0.911 | 0.968 |
| AF | 3,407 | 3,409 | 1.306 | 0.192 | 0.600 |
| BD | 3,407 | 3,409 | 0.561 | 0.575 | 0.931 |
| BMI | 3,407 | 3,409 | -0.976 | 0.329 | 0.730 |
| CRC | 3,407 | 3,409 | 0.984 | 0.325 | 0.730 |
| BC | 3,407 | 3,409 | -0.995 | 0.320 | 0.730 |
| CAL | 3,407 | 3,409 | -1.786 | 0.074 | 0.326 |
| CVD | 3,407 | 3,409 | -0.009 | 0.993 | 0.993 |
| CED | 3,407 | 3,409 | 1.280 | 0.200 | 0.600 |
| CAD | 3,407 | 3,409 | 0.231 | 0.818 | 0.961 |
| DOA | 3,407 | 3,409 | -0.326 | 0.745 | 0.961 |
| EOC | 3,407 | 3,409 | -2.167 | 0.030 | 0.155 |
| EBMDT | 3,407 | 3,409 | -6.111 | 1.04E-09 | **2.65E-08** |
| EGCR | 3,407 | 3,409 | 0.413 | 0.680 | 0.961 |
| EGCY | 3,407 | 3,409 | -0.210 | 0.834 | 0.961 |
| HBA1C_DF | 3,407 | 3,409 | 0.130 | 0.896 | 0.968 |
| HEIGHT | 3,407 | 3,409 | 4.351 | 1.38E-05 | **2.35E-04** |
| HDL | 3,407 | 3,409 | 0.294 | 0.769 | 0.961 |
| HT | 3,407 | 3,409 | -0.884 | 0.377 | 0.743 |
| IOP | 3,407 | 3,409 | -2.366 | 0.018 | 0.151 |
| ISS | 3,407 | 3,409 | 0.066 | 0.947 | 0.986 |
| LDL_SF | 3,407 | 3,409 | -0.193 | 0.847 | 0.961 |
| MEL | 3,407 | 3,409 | 2.659 | 0.008 | 0.080 |
| MS | 3,407 | 3,409 | -2.293 | 0.022 | 0.151 |
| OTFA | 3,407 | 3,409 | 0.318 | 0.750 | 0.961 |
| OSFA | 3,407 | 3,409 | 0.770 | 0.441 | 0.820 |
| OP | 3,407 | 3,409 | 6.484 | 9.54E-11 | **4.87E-09** |
| PD | 3,407 | 3,409 | 1.041 | 0.298 | 0.730 |
| PDCL | 3,407 | 3,409 | 0.663 | 0.507 | 0.862 |
| PHG | 3,407 | 3,409 | 0.392 | 0.695 | 0.961 |
| PFA | 3,407 | 3,409 | 0.495 | 0.621 | 0.932 |
| POAG | 3,407 | 3,409 | -1.083 | 0.279 | 0.730 |
| PC | 3,407 | 3,409 | 0.675 | 0.500 | 0.862 |
| PSO | 3,407 | 3,409 | -2.234 | 0.026 | 0.151 |
| RMNC | 3,407 | 3,409 | 0.501 | 0.617 | 0.932 |

| | | | | | |
|---|---|---|---|---|---|
| RHR | 3,407 | 3,409 | -2.865 | 0.004 | 0.053 |
| RA | 3,407 | 3,409 | 0.297 | 0.766 | 0.961 |
| SCZ | 3,407 | 3,409 | -0.880 | 0.379 | 0.743 |
| SGM | 3,407 | 3,409 | -0.224 | 0.823 | 0.961 |
| SLE | 3,407 | 3,409 | 1.458 | 0.145 | 0.493 |
| TCH | 3,407 | 3,409 | 0.191 | 0.848 | 0.961 |
| TFA | 3,407 | 3,409 | 0.892 | 0.372 | 0.743 |
| TTG | 3,407 | 3,409 | 1.212 | 0.226 | 0.640 |
| T1D | 3,407 | 3,409 | 1.771 | 0.077 | 0.326 |
| T2D | 3,407 | 3,409 | -2.218 | 0.027 | 0.151 |
| VTE | 3,407 | 3,409 | -1.613 | 0.107 | 0.390 |

**Appendix 1—table 11. Most significant single-variant associations ($P < 5 \times 10^{-8}$) detected in the GWAS analyses.** Six independent SNPs at genome-wide significance level were identified by linkage disequilibrium (LD) clumping ($r2 < 0.1$ within a 250 kb window). The location (chromosome [chr] and base position [bp]), alleles (A1 = effect allele and A2 = other allele), effect (β) and its standard error (β SE) with respect to A1, and association p-values from regression model of the variants are given, along with functional consequences of SNPs on gene by performing ANNOVAR.

| SNP | A1 | A2 | p-value | β | β SE | Location (chr:bp) | Gene symbol | Position relative to gene |
|---|---|---|---|---|---|---|---|---|
| rs10835187 | C | T | 1.70e-14 | -0.02558 | 0.003333 | 11:27505677 | LGR4, LIN7C | intergenic |
| rs7776725 | C | T | 4.47e-13 | -0.02640 | 0.003644 | 7:121033121 | FAM3C | intronic |
| rs779233904 | AAC | A | 1.57e-09 | 0.02083 | 0.003449 | 6:151910404 | CCDC170 | intronic |
| rs2504071 | T | C | 3.34e-09 | 0.01959 | 0.003311 | 6:152084862 | ESR1 | intronic |
| 17:43553496:A:AAT | A | AAT | 6.65e-09 | -0.02472 | 0.004261 | 17:43553496 | PLEKHM1 | intronic |
| 10:104227791:G:GA | GA | G | 1.48e-08 | -0.01889 | 0.003334 | 10:104227791 | TMEM180 | intronic |

**Appendix 1—table 12. Association between gene expression profiles of mapped genes and estimated APC during brain development.** The bold P values reflect significance after the spatial permutation test.

| **Gene Symbol** | **Spearman's ρ** | **Pvalue** | **P.permutation** |
|---|---|---|---|
| ACTR1A | 0.096 | 0.440 | 0.328 |
| ARHGAP27 | 0.051 | 0.684 | 0.445 |
| ARL17B | 0.047 | 0.705 | 0.452 |
| BDNF-AS | 0.256 | 0.038 | **0.038** |
| CCDC170 | 0.268 | 0.030 | 0.069 |
| ESR1 | 0.021 | 0.870 | 0.483 |
| FAM3C | -0.096 | 0.444 | 0.356 |
| KANSL1 | -0.262 | 0.034 | 0.073 |
| KANSL1-AS1 | 0.067 | 0.594 | 0.313 |
| LGR4 | 0.558 | 1.78E-06 | **2.50E-04** |
| LIN7C | 0.036 | 0.775 | 0.464 |
| LRRC37A4P | -0.272 | 0.027 | 0.148 |
| MAPT | 0.024 | 0.846 | 0.405 |
| PLEKHM1 | -0.276 | 0.025 | 0.109 |
| SPPL2C | 0.116 | 0.351 | 0.189 |
| STH | -0.147 | 0.238 | 0.277 |
| SUFU | 0.407 | 0.001 | **0.028** |

**Appendix 1—table 13. Association between gene expression profiles of mapped genes and estimated APC during brain aging.** The bold P values reflect significance after the spatial permutation test.

| **Gene Symbol** | **Spearman's ρ** | **Pvalue** | **P.permutation** |
|---|---|---|---|
| ACTR1A | -0.235 | 0.058 | 0.052 |
| ARHGAP27 | 0.486 | 4.48E-05 | **5.50E-04** |
| ARL17B | 0.090 | 0.473 | 0.240 |
| BDNF-AS | 0.490 | 3.68E-05 | **1.50E-04** |
| CCDC170 | 0.206 | 0.098 | 0.075 |
| ESR1 | 0.532 | 6.02E-06 | **1.50E-04** |
| FAM3C | -0.366 | 0.003 | **0.005** |
| KANSL1 | 0.213 | 0.086 | 0.078 |
| KANSL1-AS1 | -0.262 | 0.034 | 0.059 |
| LGR4 | 0.070 | 0.576 | 0.348 |
| LIN7C | 0.177 | 0.154 | 0.120 |
| LRRC37A4P | 0.143 | 0.250 | 0.165 |
| MAPT | -0.287 | 0.020 | **0.022** |
| PLEKHM1 | 0.202 | 0.104 | 0.080 |
| SPPL2C | 0.211 | 0.089 | 0.059 |
| STH | -0.001 | 0.997 | 0.490 |
| SUFU | -0.036 | 0.773 | 0.373 |

**Appendix 1—table 14. Model evaluation results using relative measures: AIC, BIC, likelihood ratio test and intra-class correlation (ICC).**

| model | AIC | BIC | lrtest | ICC_adjusted | ICC_unadjusted |
|---|---|---|---|---|---|
| lmer_intr_thalamus.proper | 79444.24 | 79591.29 | 6.45E-15 | 0.8875 | 0.3958 |
| lmer_slope_thalamus.proper | 79382.89 | 79547.24 | 6.45E-15 | 0.8889 | 0.3986 |
| lmer_intr_caudate | 95845.48 | 95992.54 | 2.68E-71 | 0.9543 | 0.6824 |
| lmer_slope_caudate | 95524.49 | 95688.84 | 2.68E-71 | 0.9564 | 0.6817 |
| lmer_intr_putamen | 92106.06 | 92253.12 | 2.40E-42 | 0.9398 | 0.5946 |
| lmer_slope_putamen | 91918.4 | 92082.75 | 2.40E-42 | 0.9424 | 0.5933 |
| lmer_intr_pallidum | 90500.3 | 90647.36 | 4.21E-42 | 0.8859 | 0.5116 |
| lmer_slope_pallidum | 90313.76 | 90478.12 | 4.21E-42 | 0.8862 | 0.5093 |
| lmer_intr_hippocampus | 89833.92 | 89980.97 | 1.37E-08 | 0.9309 | 0.5505 |
| lmer_slope_hippocampus | 89801.71 | 89966.06 | 1.37E-08 | 0.9329 | 0.5505 |
| lmer_intr_amygdala | 89976.98 | 90124.03 | 4.01E-06 | 0.8697 | 0.4898 |
| lmer_slope_amygdala | 89956.13 | 90120.48 | 4.01E-06 | 0.8713 | 0.4897 |
| lmer_intr_accumbens.area | 96936.19 | 97083.24 | 1.69E-14 | 0.8228 | 0.5295 |
| lmer_slope_accumbens.area | 96876.77 | 97041.12 | 1.69E-14 | 0.8233 | 0.5310 |
| lmer_intr_bankssts | 97981.96 | 98129.01 | 0.001818 | 0.9339 | 0.6798 |
| lmer_slope_bankssts | 97973.34 | 98137.69 | 0.001818 | 0.9354 | 0.6814 |
| lmer_intr_caudal.anterior.cingulate | 109133.2 | 109280.3 | 0.131507 | 0.8638 | 0.7653 |
| lmer_slope_caudal.anterior.cingulate | 109133.2 | 109297.5 | 0.131507 | 0.8644 | 0.7659 |
| lmer_intr_caudal.middle.frontal | 93118.13 | 93265.19 | 8.31E-06 | 0.9227 | 0.5929 |
| lmer_slope_caudal.middle.frontal | 93098.74 | 93263.09 | 8.31E-06 | 0.9246 | 0.5949 |
| lmer_intr_cuneus | 101801.4 | 101948.5 | 0.014429 | 0.9242 | 0.7256 |
| lmer_slope_cuneus | 101796.9 | 101961.3 | 0.014429 | 0.9245 | 0.7262 |
| lmer_intr_entorhinal | 109404.9 | 109551.9 | 1.63E-14 | 0.8013 | 0.6908 |
| lmer_slope_entorhinal | 109345.4 | 109509.7 | 1.63E-14 | 0.8037 | 0.6928 |
| lmer_intr_fusiform | 87545.88 | 87692.93 | 9.17E-05 | 0.9139 | 0.5062 |
| lmer_slope_fusiform | 87531.28 | 87695.63 | 9.17E-05 | 0.9163 | 0.5074 |
| lmer_intr_inferior.parietal | 89044.43 | 89191.48 | 5.89E-14 | 0.9374 | 0.5564 |
| lmer_slope_inferior.parietal | 88987.51 | 89151.86 | 5.89E-14 | 0.9419 | 0.5603 |
| lmer_intr_inferior.temporal | 84066.47 | 84213.52 | 5.73E-07 | 0.9384 | 0.4956 |
| lmer_slope_inferior.temporal | 84041.73 | 84206.08 | 5.73E-07 | 0.9405 | 0.4977 |
| lmer_intr_isthmus.cingulate | 92442.12 | 92589.17 | 0.127191 | 0.9275 | 0.5862 |
| lmer_slope_isthmus.cingulate | 92442 | 92606.35 | 0.127191 | 0.9284 | 0.5869 |
| lmer_intr_lateral.occipital | 89550.4 | 89697.45 | 0.003943 | 0.9121 | 0.5273 |
| lmer_slope_lateral.occipital | 89543.33 | 89707.68 | 0.003943 | 0.9129 | 0.5282 |
| lmer_intr_lateral.orbitofrontal | 86224.13 | 86371.18 | 1.29E-08 | 0.8466 | 0.4323 |
| lmer_slope_lateral.orbitofrontal | 86191.79 | 86356.14 | 1.29E-08 | 0.8497 | 0.4345 |
| lmer_intr_lingual | 102605.2 | 102752.2 | 0.041242 | 0.9181 | 0.7268 |
| lmer_slope_lingual | 102602.8 | 102767.2 | 0.041242 | 0.9181 | 0.7272 |

| | | | | | |
|---|---|---|---|---|---|
| lmer_intr_medial.orbitofrontal | 89954.36 | 90101.41 | 2.25E-06 | 0.7832 | 0.4219 |
| lmer_slope_medial.orbitofrontal | 89932.35 | 90096.7 | 2.25E-06 | 0.7872 | 0.4241 |
| lmer_intr_middle.temporal | 83331.01 | 83478.06 | 0.0019 | 0.9177 | 0.4615 |
| lmer_slope_middle.temporal | 83322.48 | 83486.83 | 0.0019 | 0.9195 | 0.4629 |
| lmer_intr_parahippocampal | 108997.1 | 109144.2 | 4.99E-05 | 0.8686 | 0.7639 |
| lmer_slope_parahippocampal | 108981.3 | 109145.6 | 4.99E-05 | 0.8690 | 0.7639 |
| lmer_intr_paracentral | 98058.63 | 98205.68 | 0.015686 | 0.8695 | 0.5958 |
| lmer_slope_paracentral | 98054.32 | 98218.67 | 0.015686 | 0.8705 | 0.5960 |
| lmer_intr_pars.opercularis | 96829.05 | 96976.1 | 1.20E-12 | 0.9354 | 0.6658 |
| lmer_slope_pars.opercularis | 96778.15 | 96942.5 | 1.20E-12 | 0.9396 | 0.6698 |
| lmer_intr_pars.orbitalis | 96989.61 | 97136.67 | 1.39E-05 | 0.8785 | 0.5892 |
| lmer_slope_pars.orbitalis | 96971.25 | 97135.6 | 1.39E-05 | 0.8805 | 0.5912 |
| lmer_intr_pars.triangularis | 96637.58 | 96784.63 | 4.55E-18 | 0.9402 | 0.6710 |
| lmer_slope_pars.triangularis | 96561.72 | 96726.07 | 4.55E-18 | 0.9439 | 0.6748 |
| lmer_intr_pericalcarine | 105115.9 | 105263 | 0.022841 | 0.9429 | 0.8190 |
| lmer_slope_pericalcarine | 105112.4 | 105276.7 | 0.022841 | 0.9441 | 0.8200 |
| lmer_intr_postcentral | 91605.6 | 91752.65 | 0.000549 | 0.8764 | 0.5189 |
| lmer_slope_postcentral | 91594.59 | 91758.94 | 0.000549 | 0.8789 | 0.5203 |
| lmer_intr_posterior.cingulate | 96853.73 | 97000.78 | 2.21E-19 | 0.8913 | 0.6014 |
| lmer_slope_posterior.cingulate | 96771.82 | 96936.17 | 2.21E-19 | 0.8949 | 0.6022 |
| lmer_intr_precentral | 91186.59 | 91333.64 | 2.61E-12 | 0.8478 | 0.4864 |
| lmer_slope_precentral | 91137.25 | 91301.6 | 2.61E-12 | 0.8504 | 0.4864 |
| lmer_intr_precuneus | 84734.58 | 84881.63 | 1.23E-08 | 0.9088 | 0.4708 |
| lmer_slope_precuneus | 84702.16 | 84866.51 | 1.23E-08 | 0.9126 | 0.4730 |
| lmer_intr_rostral.anterior.cingulate | 95688.68 | 95835.73 | 2.64E-12 | 0.9093 | 0.6097 |
| lmer_slope_rostral.anterior.cingulate | 95639.36 | 95803.72 | 2.64E-12 | 0.9122 | 0.6129 |
| lmer_intr_rostral.middle.frontal | 80873.84 | 81020.89 | 2.57E-17 | 0.9137 | 0.4354 |
| lmer_slope_rostral.middle.frontal | 80801.44 | 80965.79 | 2.57E-17 | 0.9191 | 0.4395 |
| lmer_intr_superior.frontal | 79730.6 | 79877.65 | 1.25E-11 | 0.8921 | 0.4038 |
| lmer_slope_superior.frontal | 79684.38 | 79848.73 | 1.25E-11 | 0.8972 | 0.4060 |
| lmer_intr_superior.parietal | 92895.95 | 93043 | 8.55E-07 | 0.8899 | 0.5487 |
| lmer_slope_superior.parietal | 92872.01 | 93036.36 | 8.55E-07 | 0.8934 | 0.5512 |
| lmer_intr_superior.temporal | 86495.24 | 86642.29 | 0.003021 | 0.9148 | 0.4959 |
| lmer_slope_superior.temporal | 86487.64 | 86651.99 | 0.003021 | 0.9168 | 0.4971 |
| lmer_intr_supramarginal | 87390.39 | 87537.44 | 3.67E-13 | 0.9263 | 0.5221 |
| lmer_slope_supramarginal | 87337.12 | 87501.47 | 3.67E-13 | 0.9320 | 0.5258 |
| lmer_intr_frontal.pole | 110426.3 | 110573.4 | 1.95E-65 | 0.6907 | 0.5868 |
| lmer_slope_frontal.pole | 110132.3 | 110296.7 | 1.95E-65 | 0.6957 | 0.5882 |
| lmer_intr_transverse.temporal | 102753.2 | 102900.2 | 0.032413 | 0.9130 | 0.7247 |
| lmer_slope_transverse.temporal | 102750.3 | 102914.7 | 0.032413 | 0.9144 | 0.7258 |
| lmer_intr_insula | 88693.2 | 88840.26 | 7.61E-14 | 0.8263 | 0.4416 |
| lmer_slope_insula | 88636.79 | 88801.14 | 7.61E-14 | 0.8290 | 0.4420 |

#### Appendix 1: Supplementary Figures

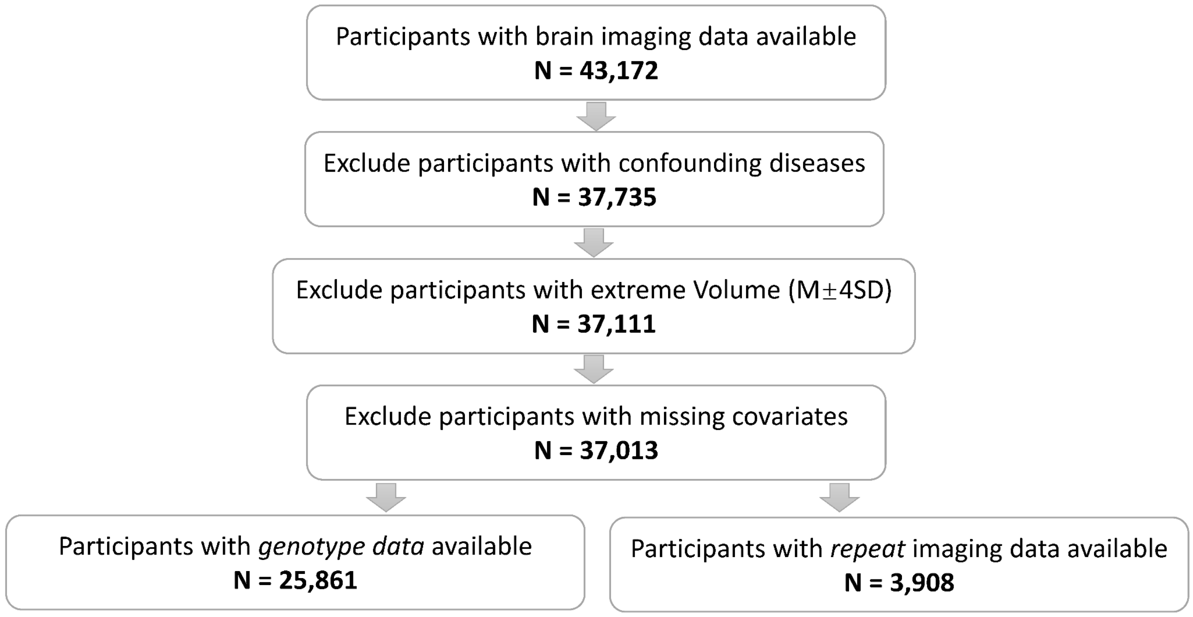


**Appendix 1—figure 1. The sample selection workflow.** Confounding diseases that resulted in participant exclusion were listed in Appendix 1—table 1 and S2. Covariates that were required for inclusion were age, sex, assessment center, handedness and ethnicity. Participants included in neurodevelopmental and neurodegenerative disorders PRS and brain aging pattern GWAS analysis were of 'White-British' background and passed all other QC measures. When describing the mirroring pattern of the brain, people who have repeated imaging measurements are included.

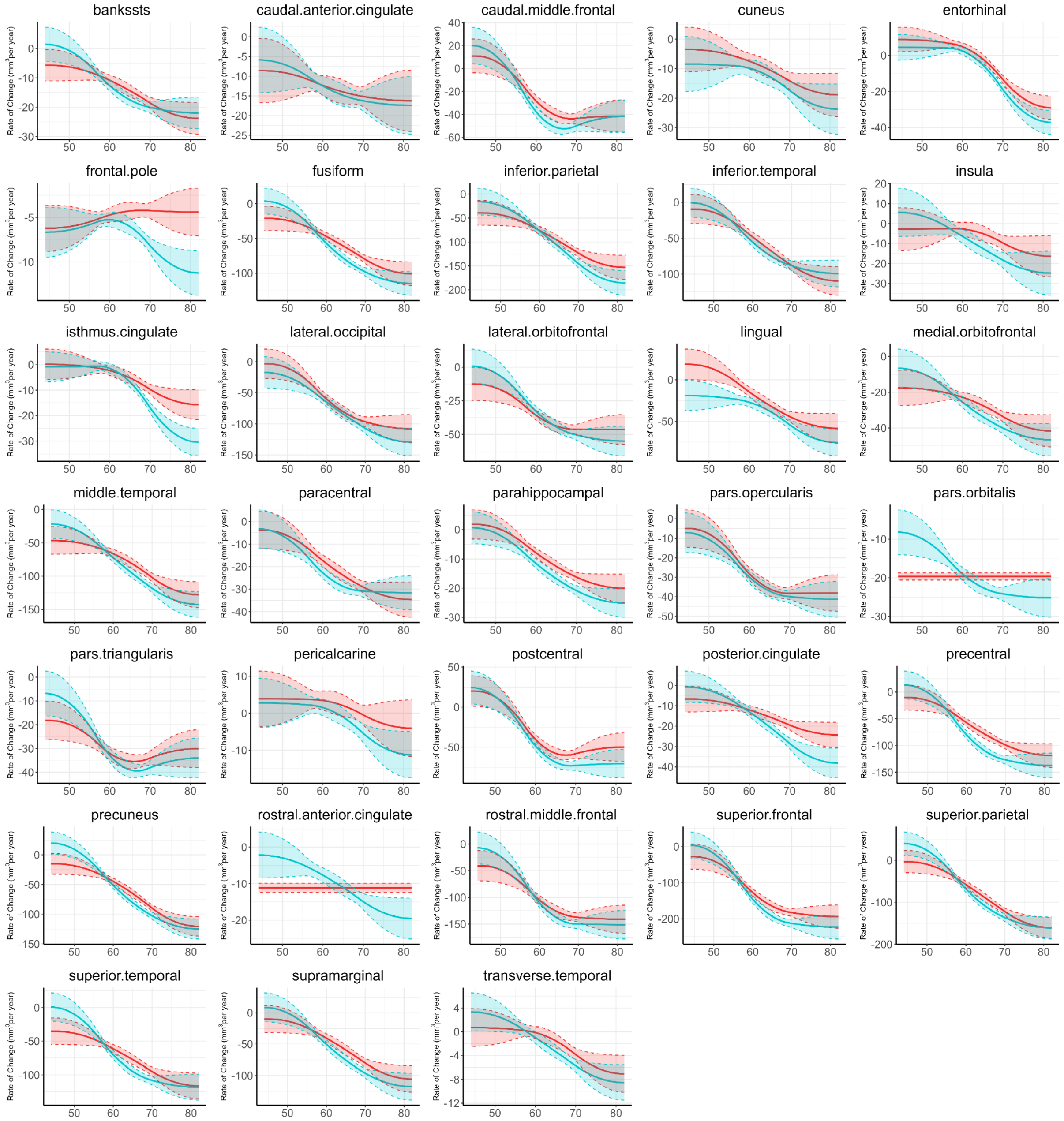


**Appendix 1—figure 2. Estimated rates of change in regional volumes for 33 bilateral brain regions.** The shaded area around the fit line denotes the 95% CI. Rates of volumetric change for two aging patterns were estimated by the first derivatives of the median whole-brain standardized trajectories. Whole-brain standardized gray matter volume trajectories in mid-to-late adulthood were estimated for two patterns using GAMM, incorporating both cross-sectional and longitudinal information from a total of 40,921 observations. Covariates including sex, site, handedness, ethnic, and ICV were considered in the analysis.

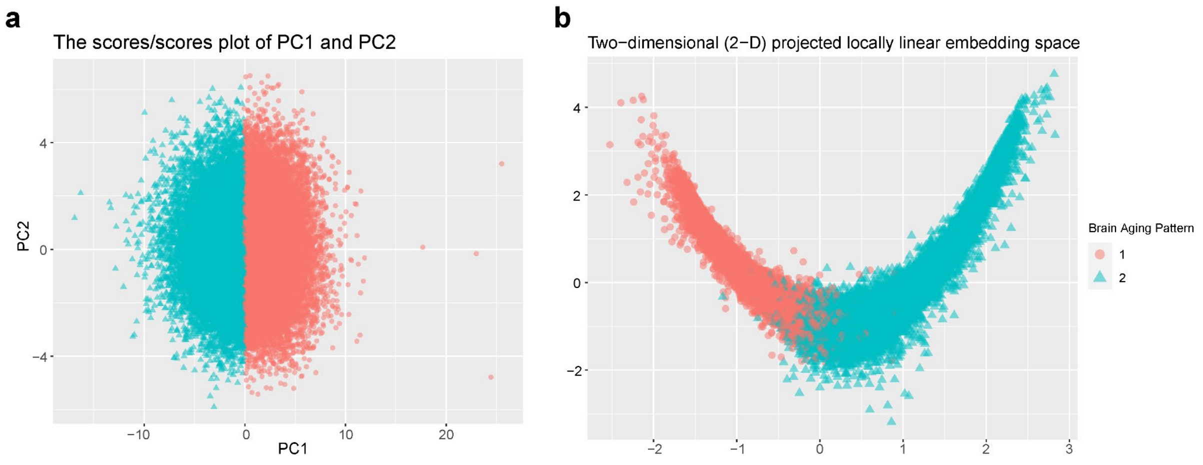


**Appendix 1—figure 3. Stratification of the identified brain aging patterns using linear and non-linear dimensionality reduction methods.** (a) The principal component space of PC1 and PC2, and (b) two-dimensional projected locally linear embedding space derived from brain volumetric measures. Points have been colored and shaped according to grouping labels of the brain aging patterns.

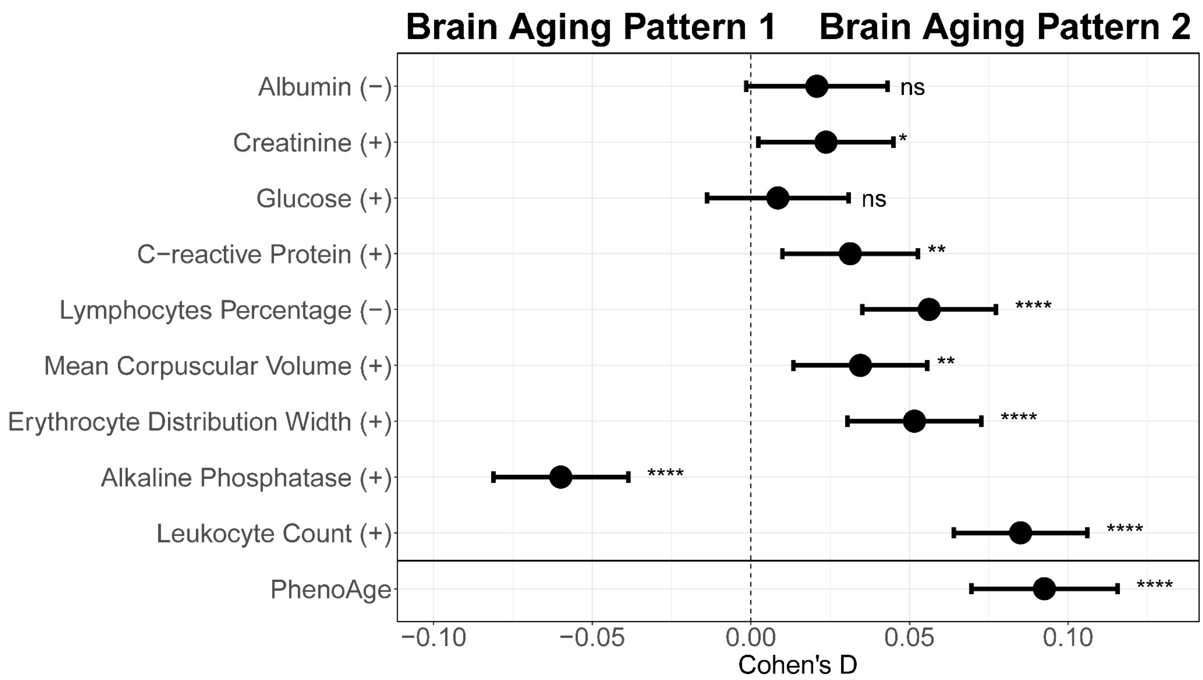


**Appendix 1—figure 4. Effect size for comparing each individual blood biochemical metric (used to calculate the PhenoAge) between participants with brain aging patterns 1 and 2.** Results were presented such that each blood biochemical metric is in the same direction as their weights in calculating PhenoAge (indicated by the sign in parentheses after the metric name). Positive Cohen's D indicate older biological age in participants with brain aging pattern 2 compared to those with pattern 1. Width of the lines extending from the center point represent 95% confidence interval. Two-sided P values were obtained using adjusted (for sex, age, ethnic, BMI, smoking status, alcohol frequency and education years) multivariate regression models. Stars indicate statistical significance after FDR correction for 9 comparisons. ****: p <= 0.0001, **: p <= 0.01, *: p <= 0.05, ns: p > 0.05.

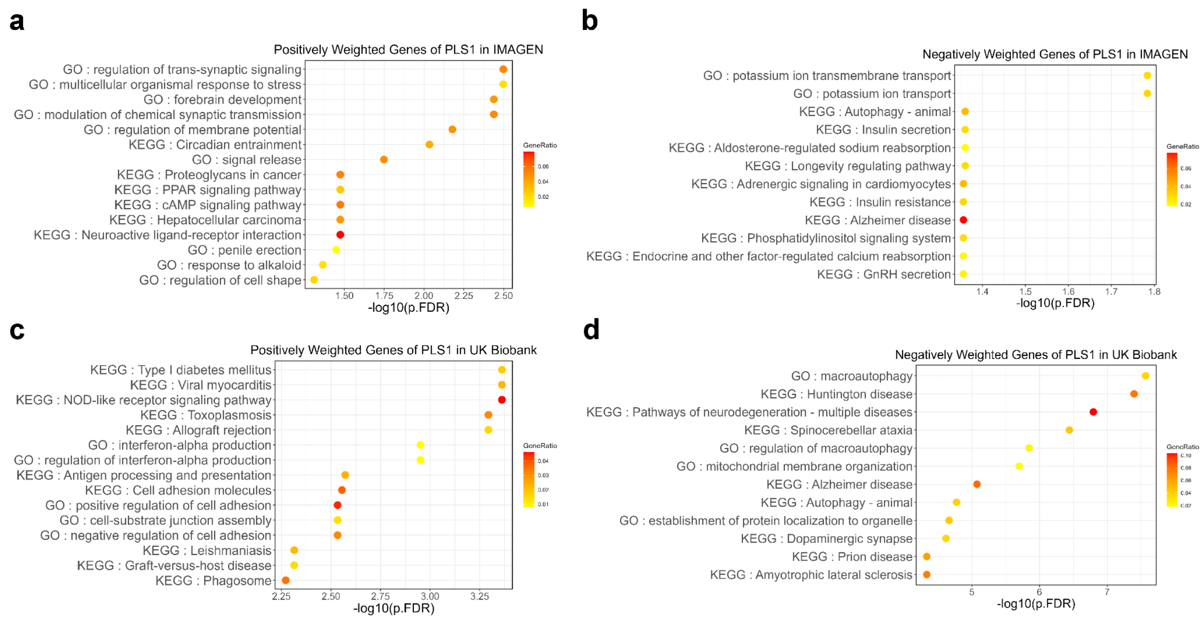


**Appendix 1—figure 5. Gene set enrichment of Kyoto Encyclopedia of Genes and Genomes (KEGG) pathways and gene ontology (GO) of biological processes.** 990 genes (a) or 1,149 genes (b) were spatially positively or negatively correlated with the first PLS component of delayed structural brain development, and 2,081 genes (c) or 2,293 genes (d) were spatially positively or negatively correlated with the first PLS component of accelerated structural brain aging.

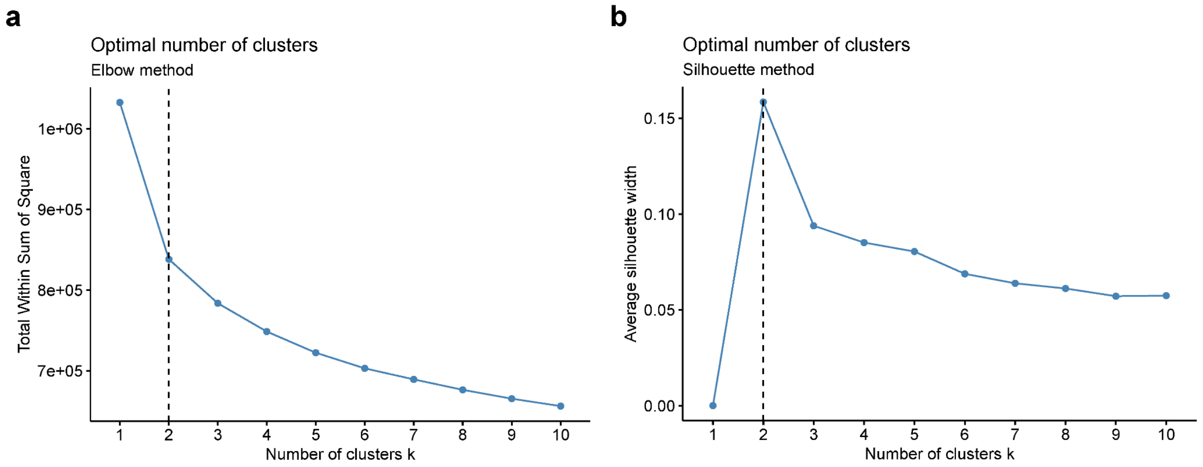


**Appendix 1—figure 6. Optimal number of clusters was chosen using elbow method (a) and silhouette method (b).** Both elbow method and silhouette method suggest that 2 should be determined as the optimal number of clusters.
